# Curricular change in a medical school: a new method for analysis of students’ academic pathways

**DOI:** 10.1101/2020.04.25.20079715

**Authors:** Damián Canales Sánchez, Tomás Bautista Godínez, J. Gerardo Moreno Salinas, Manuel García-Minjares, Melchor Sánchez-Mendiola

## Abstract

**Background:** Curricular changes in medical schools occur due to advances in medical sciences, but its evaluation is limited and fragmented, with scant data of students’ success as they progress through the program. Longitudinal follow-up can be “natural experiments” to explore innovative methodologies.

**Objective:** To propose a method for analyzing students’ academic pathways, and identify changes associated with a medical school curricular reform.

**Methods:** We analyzed the academic pathways of students throughout the program in two different curricula (1993 and 2010), at the National Autonomous University of Mexico (UNAM) Faculty of Medicine. Advancement of each student in the program was calculated with the “academic trajectory” using the accumulated credits in a defined time period, and the percentage of students that completed the credits for each stage of the curriculum. The theoretical framework is based on the “life course” approach, applying concepts of trajectory, transition and state. Data was analyzed with R software and TraMineR algorithm.

**Results:** Five student cohorts of the older curriculum were studied (classes of 1994, 1995, 1996, 2004 and 2005), and two cohorts of the new curriculum (2011, 2012), a population of 6829 students. Students in the newer curriculum had a faster, more timely and efficient advancement in academic pathways, than cohorts in the older one. There was a higher percentage of students with “regular” trajectory (without failed courses) in the newer curriculum. Regularity is a straightforward metric that allows identification of complex curricular changes’ associated effects.

**Conclusions:** Analysis of students’ academic pathways offers valuable information to evaluate curricular changes, which is difficult to obtain with traditional cross-sectional studies. The study does not provide proof of causality regarding the educational impact of different programs, although it can be useful to complement the array of program evaluation strategies in higher education.

## Introduction

Higher education throughout the world is facing several complex challenges, one of the more difficult is the evaluation of curricular changes [1,2]. Even though universities spend a great amount of resources planning and implementing educational programs, a major issue is the intricate problem of evaluating the impact of curricula in students’ academic performance [3–5]. It is not straightforward to identify the individual components of a major curricular change and their effects on the plethora of outcome variables, indicators of teacher performance and student learning, since there are many interlinked variables that can influence the main outcomes [3,4,6].

There are few studies that explore the influence of the curriculum in students’ performance during and after long-term formal educational interventions in higher education, probably due to the following: curricular changes are influenced by many contextual political, economic, and teachers’ idiosyncratic ways of conceptualizing the educational process, which creates a difficult scenario for experimental research designs, like controlled educational trials; universities are frequently reluctant to publish the results of their educational outcome variables, and school authorities are cautious to display the results of curricular changes, particularly when those results are negative; there is not much institutional motivation to publish the impact of curricular changes, since program modifications are the result of a lengthy political process, and when change has been implemented it is difficult, if not impossible, to return to the previous curricular state; and finally, curricular longitudinal analyses or pre-post studies are likely to be questioned about their internal validity, since there are several validity threats that limit causal inferences in this type of research design [3,4,7–10].

Medical schools’ instruction is of major social importance because trainees and graduates apply their knowledge and abilities in human beings, sometimes in life and death situations [11–13]. In the past century, there have been many major proposals for radically changing medical education and medical schools’ curricula, which have been met by varying degrees of resistance across the globe [14–16]. Curricular change is one of the single most difficult academic, political and educational challenges of any dean or institutional governing body, because it implies a huge effort by all involved, and changes are quite difficult to sustain and measure [7,17]. Even though all medical schools, old and new, undergo curricular change in a semi-permanent fashion, the publication of these experiences are usually case reports, and frequently they are focused on the description of an educational model (competency based, integrated curriculum, problem-based learning), faculty development strategies, organizational change and leadership challenges, particular aspects of teaching a specific discipline (biomedical informatics), or an individual competency (e.g. communication skills, professionalism, empathy and compassion) [7,18–20].

There is a dearth of publications related to the impact or longitudinal evaluation of major curricular changes, most of these related to students’ knowledge levels, their performance in summative tests, or the opinions of students and teachers regarding the curricular intervention or educational innovation [2–4,6,21–23]. The majority of curricular changes are published as books [24–26], or internal quality reports, with scant data on longitudinal follow-up, and underuse of modern methodological innovations like learning analytics and data visualization [27–31].

Terminal efficiency in higher education has been one of the main traditional indicators for universities’ accountability, and is a frequently used measure for program evaluation and accreditation [1,11,32–35]. The measurement of terminal efficiency in higher education can be done cross-sectionally or longitudinally, the first is the arithmetic ratio of the number of graduates divided by the number of students that are admitted in a specific period; the second measures terminal efficiency through life paths, the recurrent variables are of sociodemographic and family orders [32,36]. The terminal efficiency parameter is one of the main indicators used by external organizations for the accreditation of higher education programs in many countries, it is a parameter that allows quantitative identification of variables that can be influenced by curricular modifications. Curriculum changes, on the other hand, are driven by institutional policies and bylaws, and are influenced by multidimensional factors [2,18].

Traditionally, terminal efficiency rate is defined as “the proportion of students that manage to finalize each educational level, related to the total of those that initiated their studies, as many cycles before as indicated by an ideal trajectory” [37]. The National Autonomous University of Mexico (UNAM) defines terminal efficiency as “the relationship that exists between the number of students that are admitted to the university in a class/generation, and the number that graduate after they finish all the curriculum credits in a stipulated time” [38], this form of estimation has been labeled “real cohorts” [11]. The concept of “efficiency” implicitly integrates the estimation of resources, its use and what is achieved with them, including “temporality” [32]. Terminal efficiency is a referent of excellence and institutional performance, it allows evaluation based on quantitative information. Accreditation agencies consider it as an element in universities’ documentation processes, and it can be used for comparison among educational institutions [35].

Unfortunately, the terminal efficiency parameter can hide the dynamics of students’ academic advancement during their transit through the curriculum. It makes identification of students that follow atypical paths difficult, since there are students that take on larger or lesser academic loads and, in consequence, their time to completion varies. This parameter also does not consider variables that may alter the students’ academic trajectories [39,40]. Longitudinal studies, as opposed to cross-sectional estimations, offer evidence of what happens between two extreme points in a real cohort [11]. Under this perspective, follow-up of students is considered a method of longitudinal analysis of academic trajectories, based on the reconstruction of the path followed by student cohorts from their entry to the cycle until its end (via dropout or promotion) [34]. An academic trajectory is the journey followed by a cohort of students in a determined time period, after its entry to a specific program of studies [38]. These studies allow the quantification of students’ academic trajectories during its path through admission, continuity and graduation, until the students fulfill the requirements of the curriculum [41].

Academic trajectories allow calculation and analysis of indicators that explain students’ individual and group academic behavior. Traditional indicators to measure academic trajectories are academic advancement, academic performance, terminal efficiency, academic delay and dropout, regularity and failure [35]. Academic trajectories’ studies have increased in sophistication, evolving from a descriptive function of performance indicators [42], to relational studies [43,44] and finally to play a predictive role with the incorporation of new models [45–47]. Academic trajectory studies have attempted to explain, using statistical classification models, the effect of individual, academic, institutional, economic and cultural variables that intervene in academic behavior [48]. These studies contribute to more rational decision making, in order to improve formative processes and academic administration [49,50]. There are two main challenges to the construction of academic trajectories models: the first is related to the availability of each individual’s data; the second is the appropriate ethical handling of personalized students’ information. These issues are barriers to the construction of longitudinal statistical models. The interpretation of information that is generated by more sophisticated academic trajectories’ studies, constitutes another challenge that inhibits their broader acceptance.

Academic trajectory studies have gradually achieved relevance in higher education institutions and have become an important information input in the design of institutional policy planning [49,50]. It is important to emphasize that, at least in the case of dropouts’ studies, about half of the published papers have focused on identifying and describing the students that suffer the event, and only 11% of the studies propose intervention alternatives to modify the outcome. There is also a lack of methodological rigor in the published studies in the measurement of latent variables, which decreases the validity of the results and underscores the need to use a common language in the field [48]. Another relevant aspect is the recognition that students’ situations are dynamic or can occur in different degrees, which creates another challenge for analysis [51].

In the last decade there has been an explosion of techniques that take advantage of the Big data revolution, and use learning analytics and data visualization methodologies, opening new frontiers for understanding learning and assessment in higher education. These innovative tools can help explore students’ academic trajectories during training and add a broader perspective to program evaluation and the impact of curricular changes [27,28]. Curriculum analyses are particularly appropriate to the use of these methods, since educational programs are by definition complex adaptive systems that occur in a fixed normative framework and identified time periods [29,31,52,53].

As far as we could gather, there are no published reports about evaluation of students’ academic paths/trajectories with methodologies that explore the potential effects of major curricular changes in medical schools. The main goal of this study is to present an innovative methodology to evaluate students’ academic pathways in several cohorts, before and after a major curricular reform, and show evidence of its potential usefulness.

## Materials and methods

### 2.1 Research design

Longitudinal quantitative research design, with quasi-experimental pre-post analysis comparing real cohorts of students’ academic paths/trajectories, before and after a major curricular change.

### 2.2 Setting

The National Autonomous University of Mexico (UNAM) Faculty of Medicine in Mexico City is the largest medical school in the country and one of the largest in Latin America, with almost 9,000 undergraduate students and more than 11,000 medical residents. It is a public institution and the largest producer of basic and clinical medical research in Mexico, through its affiliations with major national academic medical centers (http://www.facmed.unam.mx). Mexico has more than 170 medical schools, the curricula are heterogeneous although the majority are six-year programs (two year of basic sciences, two year of clinical rotations, one year of clinical internship and one year of mandatory social service) [54,55]. Mexican medical students are admitted to the university after a three-year bachelor phase, and their average age when entering medical school as undergraduate students is 18 years.

### 2.3 Medical school curricular change

Our medical school underwent a major curricular reform in the year 2010 (C-2010). The previous curriculum (C-1994) had a traditional focus based on the Flexnerian model and was in place for 16 years during the period 1994-2010. In 2010 there was a major curricular reform after a long and complex process of institutional reflection and academic dialogue, the program was approved in February 2010 [55]. Some characteristics of the C-2010 program are: organization by courses with a competency-based focus; definition of intermediate and outcome generic competencies; three curricular axes that articulate three areas of knowledge (biomedical, clinical, sociomedical/humanistic); four training phases with competency profiles; new courses (Biomedical Informatics, Basic-Clinical integration, Evidence-based Medicine and Clinical Epidemiology, among others); integration of the Clinical Skills Simulation Center in the curriculum with a problem-based learning methodology; a core curriculum for each course (an extensive description of the process, including the institutional diagnosis of the previous curriculum and the rationale for the changes in the newer curriculum is available in Spanish in the following link: http://www.facmed.unam.mx/_documentos/planes/mc/PEFMUNAM.pdf) [55]. The curricular change involved faculty development activities, more formative and diagnostic assessment for learning, the establishment of a Curriculum Committee for the evaluation and follow-up of the program. The curricular maps of both programs are in the S1 Appendix, and a comparative table of the main differences between them is in the S2 Appendix.

### 2.4 Sample

#### Selection of the studied cohorts

We analyzed seven cohorts/generations of the MD program. The first five cohorts underwent the C-1994 curriculum (1994, 1995, 1996, 2004, 2005), the last two cohorts had the C-2010 program (2011, 2012). The students’ distribution by year is shown in Table 1. In this context, the assumption was made that the curriculum is a foundational element that influences students’ academic paths/trajectories, since the efforts of the curriculum organization, faculty development and academic administration strategies converge in the curricular structure. The studied population including all the cohorts was composed of 6,829 students: 4,629 had the 1994 program and 2,200 the 2010 curriculum.

**Table 1.** Number of students in the MD program at UNAM Faculty of Medicine in Mexico City, by curricular cohort (n=6,829).

| **Cohort** | **Curriculum** | **Number of students** |
| --- | --- | --- |
| 1994 | C-1994 | 927 |
| 1995 | C-1994 | 891 |
| 1996 | C-1994 | 920 |
| 2004 | C-1994 | 925 |
| 2005 | C-1994 | 966 |
| 2011 | C-2010 | 1,045 |
| 2012 | C-2010 | 1,155 |

#### Selection criteria

In order to have representative samples of the cohorts, we included the first three generations of the 1994 curriculum. The initial years of a new curriculum implementation are usually accompanied by a strong organizational effort and intense faculty development and supervision activities, and in parallel there is frequently a “performance dip” due to the learning curve of the entire organization [18]. In 1999 there was a prolonged one-year strike at the university, which had a profound effect on the dynamics of curriculum delivery, so we included two generations after this major event (2004 and 2005), which was an uncontrolled disrupting variable that likely affected the students’ academic pathways. On the other hand, these two classes studied the 1994 curriculum a decade after its implementation, which provided information generated in a more mature and established curricular environment.

The newer curriculum (C-2010) samples were the first two generations that completed the program of studies in the established official time. At the time this study was performed, we only had complete data for these two cohorts.

### 2.5 Conceptual framework

We used “life course” theory as the conceptual methodological framework to analyze students’ academic trajectories. According to Elder, this theory aims to reveal the manner in which broader social forces mold the development of individual and collective life courses [56,57]. In our context, the driving forces at the university are operationalized in changes of curricular structures and programs, and it is expected that these variations promote changes in the temporal behavior of students’ trajectories.

We used the concepts of “trajectory”, “transition” and “state”. The concept of “trajectory” refers to a life or career line that can vary and change in direction, degree or proportion [58–60]. “Transition” relates to the changes in state, position or situation, that can be foreseeable or probabilistic [57,59]. State is the concept that articulates trajectories and transitions. A “state” is a situation in which the unit of analysis finds itself, in a determined moment, throughout time [59,61,62].

In our study, the basic unit of analysis is a student throughout time, and each state is defined according to the number of credits that a student achieves after a curricular year, until finishing the duration of that educational program. The concatenation of states where a student finds herself at the end of each educational cycle determines her academic path/trajectory, without taking into account whether she obtains or not the college degree.

### 2.6 Measurement methods and statistical analyses

The data utilized for the construction of the medical students’ trajectories were obtained from their academic history, the university official data source of academic advancement in credits. These were linked to the corresponding curriculum and program of studies. A credit is the value unit of an academic course or module; a curriculum or plan of studies is the combination of courses or modules (theoretical courses, laboratories, workshops, practices, seminars), assessment results and other institutional requirements which, when finished satisfactorily, provide evidence that the student finished the MD program [63].

The program of studies and credits are two variables used to establish the students’ situation after an academic year. The behavior of the academic paths/trajectories is identified through two indicators: Advancement/Progress and Regularity/Consistency. Advancement is the percentage of accumulated credits during the academic period, in relation to the requirements established in the formal curriculum. Regularity is the percentage of students that concluded satisfactorily the number of credits established in the curriculum for that year, in other words, the correspondence between the number of credits covered by a student after an academic year, and the number of credits established in the program for that time period.

We built eight different states to identify all the possible trajectories through which a student can transit throughout his/her academic career, following the same logic of the state we call regularity. Each state corresponds to a feasible possibility where a student can be situated after an academic year, according to the number of covered credits that are associated with each passed course during that period. For example, in idealized terms, a student should cover in the first year all the credits that are established in the curriculum. However, there is a possibility that the student does not pass all the required courses, and even that he doesn’t pass any of the courses required for that school year. Under this assumption we defined quintiles of academic advancement, and an “absorbent state” (C) that corresponds to total credit completion.

As seen in Table 2, a student that finished satisfactorily 100% of the required credits for an academic cycle is labeled *A*^*100*^, a student that finished between 80% and 99% of credits is labeled 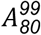, and so on and so forth until the student that did not manage to finish any credits at all. The label “A” associated with each state, represents the advancement or progress with relation to the number of credits that each student should cover in an academic cycle. The subscript in each A level means the percentage of covered credits, e.g. state 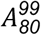 means that the student passed at least 80% of the required credits, but less than 100%. This indicator represents a measure of the student advance in his academic trajectory [36]. States have two functions: they allow the quantitative recognition of each student’s situation after each academic cycle during her academic career; and they allow comparison of the academic progress of each student in accordance with the corresponding curriculum.

**Table 2.** Interval labels of students’ transition states during their academic journey.

| States | State intervals |
| --- | --- |
| $A^{100}$ | Academic progress in agreement with the curriculum |
| $A_{80}^{99}$ | $80\% \leq \text{Academic progress} < 100\%$ |
| $A_{60}^{79}$ | $60\% \leq \text{Academic progress} < 80\%$ |
| $A_{40}^{59}$ | $40\% \leq \text{Academic progress} < 60\%$ |
| $A_{20}^{39}$ | $20\% \leq \text{Academic progress} < 40\%$ |
| $A_{10}^{19}$ | $10\% \leq \text{Academic progress} < 20\%$ |
| $A_0^9$ | $0 \leq \text{Academic progress} < 10\%$ |
| C | Completion (absorbent state) |
| NR | Not registered in the academic cycle |

#### Operationalization of states in the curricula

Course credits and the time used by a student to complete those credits allow estimation of academic progress, equal to the ratio of accumulated credits divided by the time in which credits were completed. The unit of measure of a student’s academic progress is the course credit, and passing a course is equivalent to an established number of credits in the curriculum. Accumulated credits are the sum of credits equivalent to the number of courses that a student should complete in a specified time period.

In the C-1994 curriculum, at the end of the first year six courses should be completed, for a total of 84 credits. The following year six more courses (94 credits) should be completed, for an accumulative total of 178 credits. Successively for each year, with a total of 449 credits. The sixth year is the mandatory social service that has no credit value (Table 3).

**Table 3.** Accumulated credits by year and state, UNAM’s Faculty of Medicine 1994 curriculum (C-1994)

| Year | Credits by year | Accumulated credits | State |  |  |  |  |  |  |
| --- | --- | --- | --- | --- | --- | --- | --- | --- | --- |
| | | | $A^{100}$ | $A^{99}_{80}$ | $A^{79}_{60}$ | $A^{59}_{40}$ | $A^{39}_{20}$ | $A^{19}_{10}$ | $A^9_0$ |
| 1 | 84 | 84 | 84 | 67.2 | 50.4 | 33.6 | 16.8 | 8.4 | < 8.4 |
| 2 | 94 | 178 | 178 | 142.4 | 106.8 | 71.2 | 35.6 | 17.8 | < 17.8 |
| 3 | 91 | 269 | 269 | 215.2 | 161.4 | 107.6 | 53.8 | 26.9 | < 26.9 |
| 4 | 98 | 367 | 367 | 293.6 | 220.2 | 146.8 | 73.4 | 36.7 | < 36.7 |
| 5 | 82 | 449 | 449 | 359.2 | 269.4 | 179.6 | 89.8 | 44.9 | < 44.9 |
| 6 | 0 | 449 | 449 | 359.2 | 269.4 | 179.6 | 89.8 | 44.9 | < 44.9 |

Credit accumulation begins once a student is registered for the first time in the program. This condition allows comparability of accumulated progress among students registered for different curricula, since we homologate the students’ starting point. This detail needs to be emphasized since students can be accepted to the MD program but not register for any course. Students’ accumulated credits by academic year and state can be observed in Table 3. The idealized transition state for each academic cycle is *A*^*100*^, as established in the curriculum. The extreme states are the completion state (C) and the not registered (NR) state. A student is in the C state when he accumulates the total number of credits established in the curriculum, and a student is in the NR state when he does not register for any course that year. The programs have elective courses, which the student can choose depending on his preferences. Electives have a credit designation that varies by course, and the curriculum establishes the years where electives can be taken. We established as reference the elective courses with the lower number of credits in each year.

Table 4 shows an example of students’ representative accumulative academic progress. At the end of each academic year the student is classified in one of the possible states. For example, in the first row the student passed all the courses and accumulated the total of year one credits, the same thing occurred in second (1995) and third (1996) years, so the student completed the credits in 1997.

**Table 4.** Examples of state by academic year, each row is a student of the C-1994 program (anonymized ID).

| ID | Cycle |  |  |  |  |  |  |
| --- | --- | --- | --- | --- | --- | --- | --- |
|  | 1994 | 1995 | 1996 | 1997 | 1998 | 1999 | 2000 |
| 12040296 | $A^{100}$ | $A^{100}$ | $A^{100}$ | $C$ | $C$ | $C$ | $C$ |
| 12040405 | $A_{20}^{39}$ | $A_{20}^{39}$ | $A_{40}^{59}$ | $A_{40}^{59}$ | $A_{60}^{79}$ | $A_{80}^{99}$ | $A_{80}^{99}$ |
| 12040410 | $A_{80}^{99}$ | $A_{60}^{79}$ | $A_{40}^{59}$ | $A_{20}^{39}$ | $A_{20}^{39}$ | $NR$ | $NR$ |
| 12040551 | $A_{20}^{39}$ | $A_{20}^{39}$ | $A_{40}^{59}$ | $A_{40}^{59}$ | $A_{60}^{79}$ | $A_{60}^{79}$ | $NR$ |
| 12040606 | $A_{60}^{79}$ | $A_{40}^{59}$ | $A_{40}^{59}$ | $A_{40}^{59}$ | $A_{40}^{59}$ | $A_{40}^{59}$ | $NR$ |
| 12040664 | $A_{20}^{39}$ | $A_{20}^{39}$ | $A_{40}^{59}$ | $A_{40}^{59}$ | $A_{40}^{59}$ | $A_{60}^{79}$ | $A_{80}^{99}$ |
| 12050019 | $A_{80}^{99}$ | $A_{60}^{79}$ | $A_{40}^{59}$ | $NR$ | $A_{20}^{39}$ | $A_{20}^{39}$ | $A_{20}^{39}$ |
| 12050406 | $A^{100}$ | $A^{100}$ | $A^{100}$ | $A^{100}$ | $C$ | $C$ | $C$ |
| 12050686 | $A^{100}$ | $A^{100}$ | $A^{100}$ | $A^{100}$ | $C$ | $C$ | $C$ |
| 12050929 | $A_{20}^{39}$ | $A_{20}^{39}$ | $A_{40}^{59}$ | $A_{40}^{59}$ | $A_{60}^{79}$ | $A_{60}^{79}$ | $A_{80}^{99}$ |
| 12060193 | $A_{80}^{99}$ | $A_{60}^{79}$ | $A_{40}^{59}$ | $A_{40}^{59}$ | $NR$ | $A_{40}^{59}$ | $NR$ |
| 12060824 | $A_{20}^{39}$ | $A_{20}^{39}$ | $A_{40}^{59}$ | $A_{40}^{59}$ | $A_{60}^{79}$ | $A_{80}^{99}$ | $A_{80}^{99}$ |

#### Data sources and statistical software

The main data source for the study are the academic histories of the medical students at UNAM. The area of educational assessment identified the pertinent data in the university databases, with the following fields: student identifier (anonymized for purposes of analysis), code for curricular program (C-1994 or C-2010), type of admission, courses, credits and time. The software used is R (https://www.r-project.org), specifically TraMineR “STates-Sequence” (STS). This program allowed calculation and visualization of academic trajectories [62,64].

### 2.7 Ethical aspects

The study was in compliance with the Declaration of Helsinki for research involving human subjects’ data. Data was managed anonymously in a confidential manner. The Research and Ethics Committee of UNAM Faculty of Medicine approved the research protocol as a non-invasive minimal risk study, with registration number 065/2016.

## Results

### 3.1 Descriptive statistics

The analyzed population was composed of 6,829 students, of which 4,629 (67.8%) had the 1994 curriculum and 2,200 (32.2%) the 2010 program. Table 5 shows the population descriptive statistics.

**Table 5.**
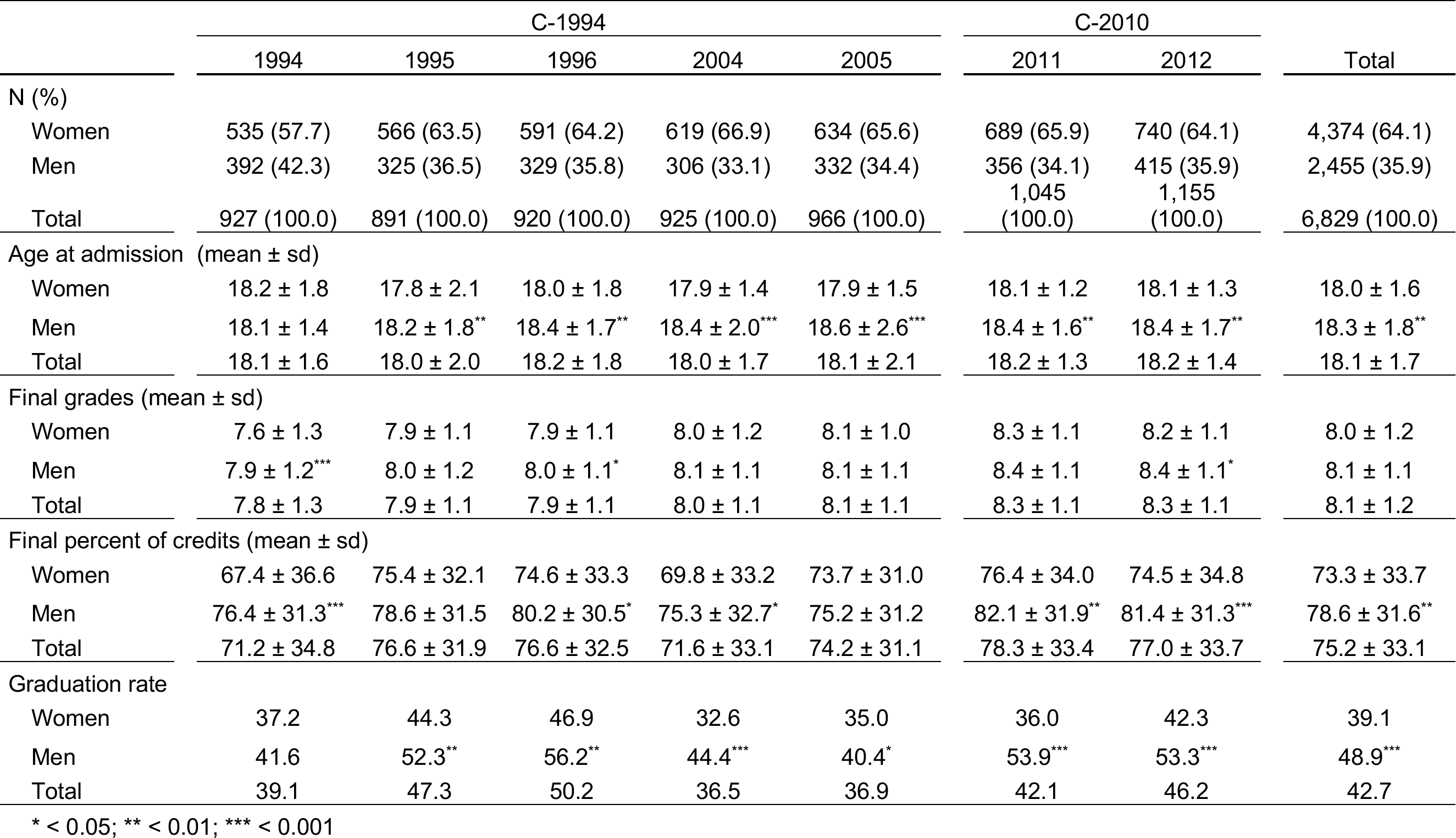
Descriptive statistics of UNAM’s Faculty of Medicine student cohorts, by program, year, gender, age at admission, final grades, final credits and graduation rates (n=6,829).

|  | C-1994 |  |  |  |  | C-2010 |  | Total |
| --- | --- | --- | --- | --- | --- | --- | --- | --- |
|  | 1994 | 1995 | 1996 | 2004 | 2005 | 2011 | 2012 |  |
| N (%) |  |  |  |  |  |  |  |  |
| Women | 535 (57.7) | 566 (63.5) | 591 (64.2) | 619 (66.9) | 634 (65.6) | 689 (65.9) | 740 (64.1) | 4,374 (64.1) |
| Men | 392 (42.3) | 325 (36.5) | 329 (35.8) | 306 (33.1) | 332 (34.4) | 356 (34.1) | 415 (35.9) | 2,455 (35.9) |
| Total | 927 (100.0) | 891 (100.0) | 920 (100.0) | 925 (100.0) | 966 (100.0) | 1,045 (100.0) | 1,155 (100.0) | 6,829 (100.0) |
| Age at admission (mean ± sd) | | | | | | | | |
| Women | 18.2 ± 1.8 | 17.8 ± 2.1 | 18.0 ± 1.8 | 17.9 ± 1.4 | 17.9 ± 1.5 | 18.1 ± 1.2 | 18.1 ± 1.3 | 18.0 ± 1.6 |
| Men | 18.1 ± 1.4 | 18.2 ± 1.8** | 18.4 ± 1.7** | 18.4 ± 2.0*** | 18.6 ± 2.6*** | 18.4 ± 1.6** | 18.4 ± 1.7** | 18.3 ± 1.8** |
| Total | 18.1 ± 1.6 | 18.0 ± 2.0 | 18.2 ± 1.8 | 18.0 ± 1.7 | 18.1 ± 2.1 | 18.2 ± 1.3 | 18.2 ± 1.4 | 18.1 ± 1.7 |
| Final grades (mean ± sd) | | | | | | | | |
| Women | 7.6 ± 1.3 | 7.9 ± 1.1 | 7.9 ± 1.1 | 8.0 ± 1.2 | 8.1 ± 1.0 | 8.3 ± 1.1 | 8.2 ± 1.1 | 8.0 ± 1.2 |
| Men | 7.9 ± 1.2*** | 8.0 ± 1.2 | 8.0 ± 1.1* | 8.1 ± 1.1 | 8.1 ± 1.1 | 8.4 ± 1.1 | 8.4 ± 1.1* | 8.1 ± 1.1 |
| Total | 7.8 ± 1.3 | 7.9 ± 1.1 | 7.9 ± 1.1 | 8.0 ± 1.1 | 8.1 ± 1.1 | 8.3 ± 1.1 | 8.3 ± 1.1 | 8.1 ± 1.2 |
| Final percent of credits (mean ± sd) | | | | | | | | |
| Women | 67.4 ± 36.6 | 75.4 ± 32.1 | 74.6 ± 33.3 | 69.8 ± 33.2 | 73.7 ± 31.0 | 76.4 ± 34.0 | 74.5 ± 34.8 | 73.3 ± 33.7 |
| Men | 76.4 ± 31.3*** | 78.6 ± 31.5 | 80.2 ± 30.5* | 75.3 ± 32.7* | 75.2 ± 31.2 | 82.1 ± 31.9** | 81.4 ± 31.3*** | 78.6 ± 31.6** |
| Total | 71.2 ± 34.8 | 76.6 ± 31.9 | 76.6 ± 32.5 | 71.6 ± 33.1 | 74.2 ± 31.1 | 78.3 ± 33.4 | 77.0 ± 33.7 | 75.2 ± 33.1 |
| Graduation rate |  |  |  |  |  |  |  |  |
| Women | 37.2 | 44.3 | 46.9 | 32.6 | 35.0 | 36.0 | 42.3 | 39.1 |
| Men | 41.6 | 52.3** | 56.2** | 44.4*** | 40.4* | 53.9*** | 53.3*** | 48.9*** |
| Total | 39.1 | 47.3 | 50.2 | 36.5 | 36.9 | 42.1 | 46.2 | 42.7 |

\* < 0.05; \*\* < 0.01; \*\*\* < 0.001

### 3.2 Transition states

The behavior of students’ academic progress for the C-1994 cohorts (1994, 1995, 1996, 2004 and 2005) is shown in Figure 1.

**Figure 1.**
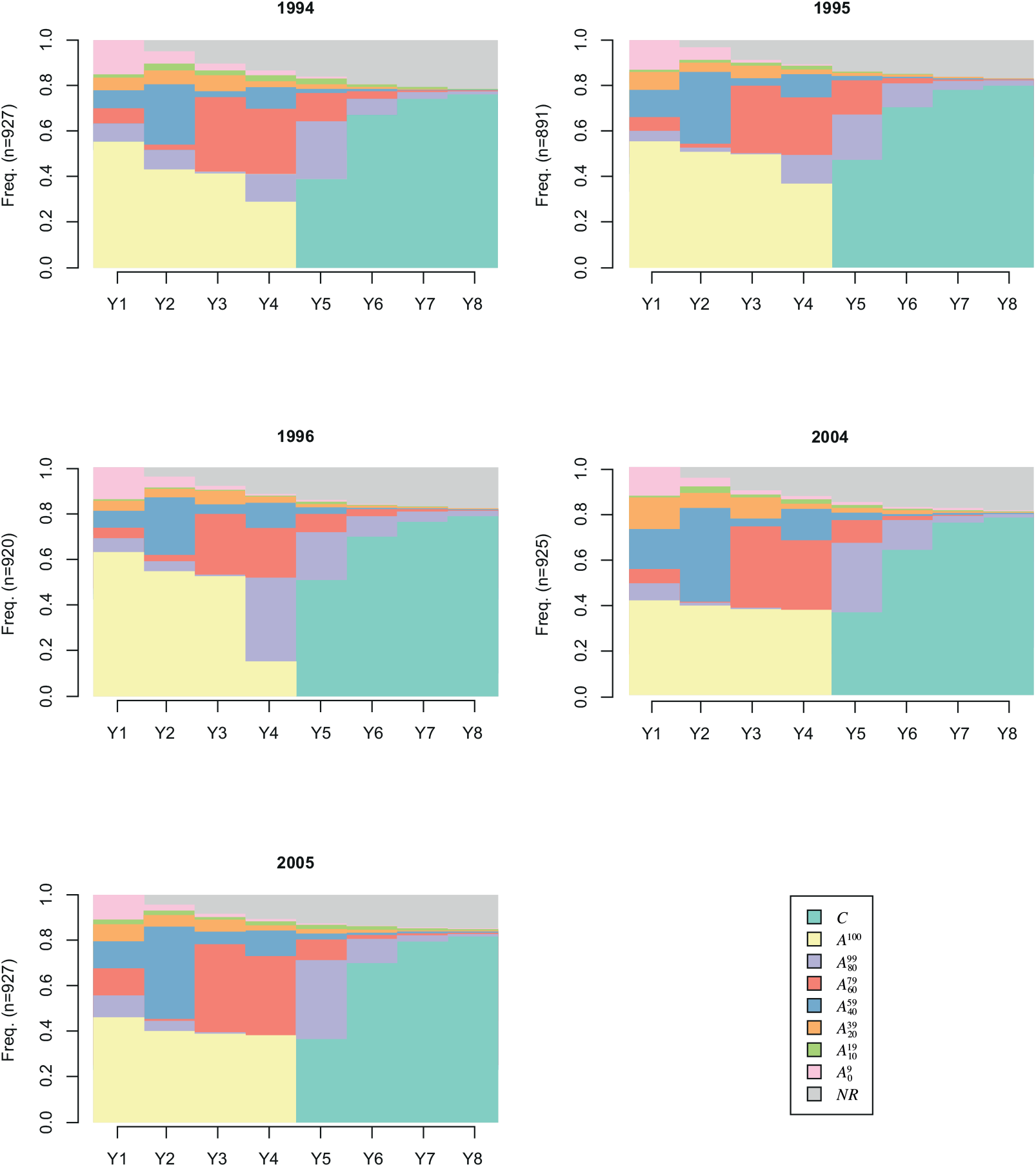
Academic progress distribution by state of five C-1994 curricular cohorts (n=4,629).

As can be seen in the advance in credits (Fig. 1), the 1994, 1995 and 1996 classes showed a similar behavior at the end of the first academic year. Six out of ten C-1994 students managed to finish in a timely manner 100% of the first year credits (*A*^*100*^). Only four of ten students from the 2004 and 2005 classes in the C-1994 curriculum finished the totality of first-year credits on time. In comparison, for the 2010 curriculum five of ten students finished 100% of the first-year credits on time (Fig. 2).

**Figure 2.**
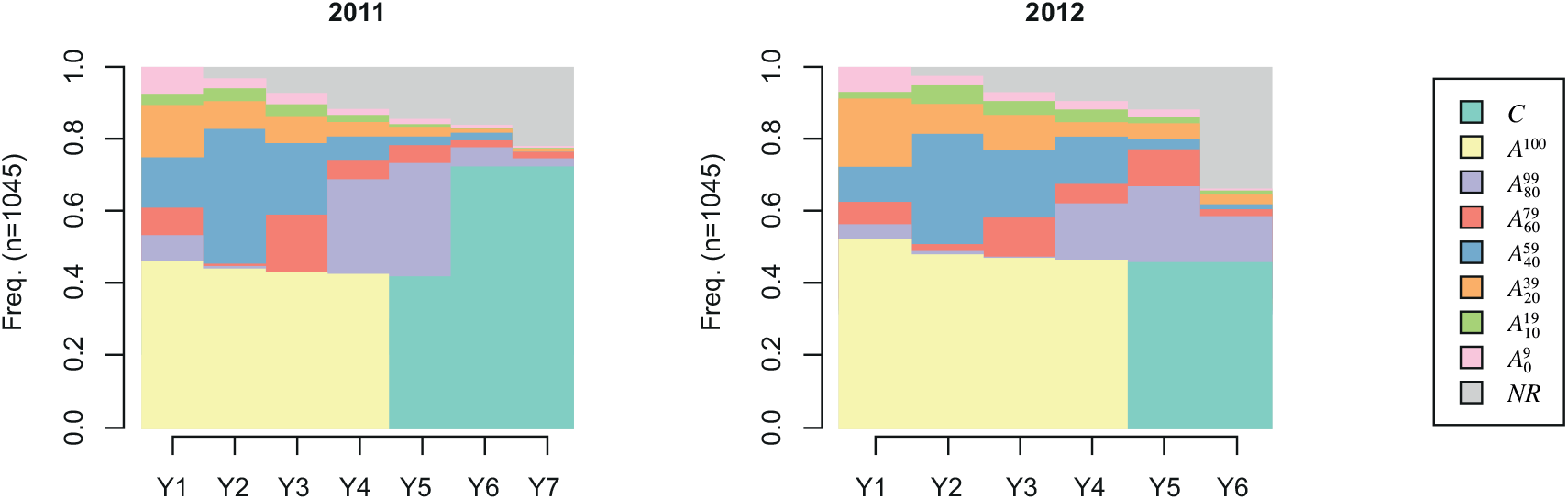
Academic progress distribution by state of two 2010 curriculum (C-2010) cohorts (n=2,200).

Despite the fact that in the earlier generations there was a larger regularity in the first year, in subsequent years the loss of regular students was accentuated, as can be seen in the gradual staggering of the 1994, 1995 and 1996 classes (Fig. 1). This could suggest that students had higher difficulties to pass the required courses. In all generations studied after 1996, regardless of the curriculum, the height of the “steps” or staggering of states decreased considerably. This behavior could indicate that the student selection mechanisms implemented after that date allowed admission of students with the needed knowledge and abilities to carry the curricular load.

By the end of the second year of the program, about 20% of the student population of the 1994, 1995 and 1996 classes covered between 40 and 59% of their academic requirements. This pattern changes in subsequent generations, in the 2011 class about 40% managed to cover the same range of academic requirements. After years three and four of the program, the 1994, 1995 and 1996 classes that had academic delay were concentrated in the 60 to 79% academic advance range, a small recovery compared to the previous year. The atypical case of the 1996 class, where only 15% of the students were regular, is likely due to the university one-year 1999 strike. In the fifth year of the program, non-regular students were mostly concentrated in the 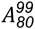 state, the remaining states are distributed first, in students that achieved 60% of academic advance, and second, in students that were in the critical state of “not registered”.

In summary, academic progress of students in generations previous to 2000 that had the C-1994 curriculum, showed a different behavior than the 2004 and 2005 classes that had the same program. In the earlier generations there was a staggered fall in regularity as students advanced in the program, reaching its lower level by the end of the fourth year. In these generations by the end of the first year, about 60% of the students were regular and at the end of the 4th year this number ranged between 20 and 40%. In the time required to cover the program’s credits, half of the students finished in a timely fashion. In generations 2004 and 2005, regularity had a constant trend of about 40%. The student group that is regular by the end of the first year maintains that state throughout the program and graduates. In both groups of generations, a year after the program stipulated time to finish the credits, a large number of students in the 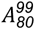 state incorporated to the graduated level, and by the eighth year, 80% of the students that were admitted to the MD program graduated.

In the C-2010 curriculum cohorts, the bands that represent the academic progress of 40 and 20% (delay of 60 and 80%) stand out throughout the curricular time period (Fig. 2). The distribution in both cohorts of C-2010 is similar. At the end of five years, the number of students that have an advance of 60% (delay of 40%) is accentuated in the 2012 cohort in the last year of studies. There is a large proportion of 80% advancement (20% delay) in the last two years of the program.

The percentage of students that have zero academic progress from the second year onwards is another behavior that stands out. In the sixth year of the 2012 cohort there is a pronounced and steep progression of students that did not register any movement. Almost two-thirds of the population find themselves in this state. In both 2011 and 2012 cohorts, the increase in students that registered zero movement is higher than any of the C-1994 cohorts.

### 3.3 Most common academic trajectories by cohort and curriculum

The 15 more frequent academic trajectories for each student population in the C-1994 curriculum are shown in Fig. 3, and those for the C-2010 program in Fig. 4. In the C-1994 cohorts, the proportion of student populations whose trajectory is shown ranges between 70.3% (cohort 1994) and 77.3% (cohort 2005). In the 2005 cohort there was a larger percentage of students that shared some of these 15 common trajectories, which was 7% lower in the 1994 cohort. The different patterns of the trajectories do not allow us to conclude that any particular cohort managed to adapt or not to the curriculum.

**Figure 3.**
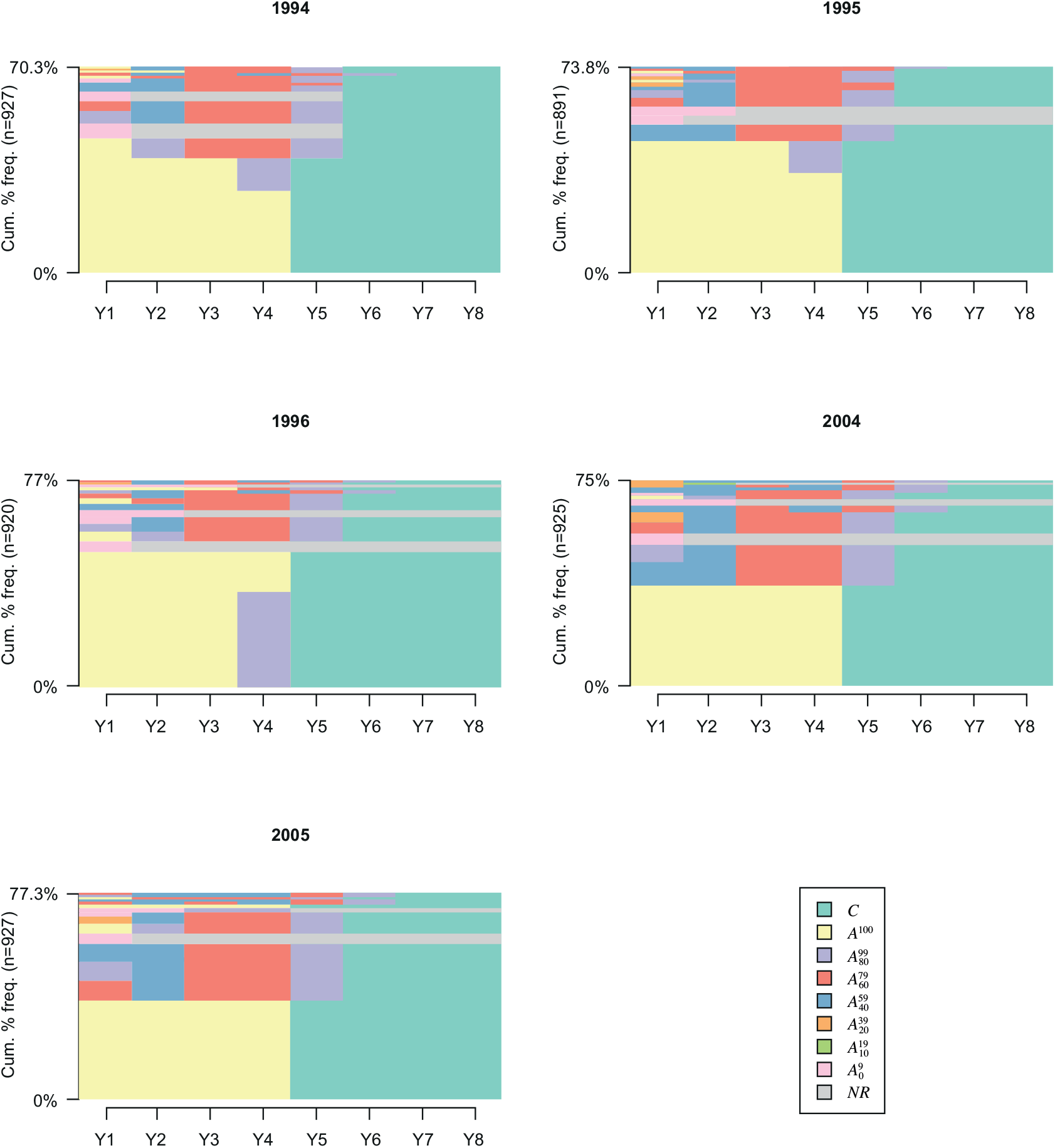
Frequency of academic progress trajectories in the C-1994 program 1994, 1995, 2004 student cohorts, UNAM Faculty of Medicine.

**Figure 4.**
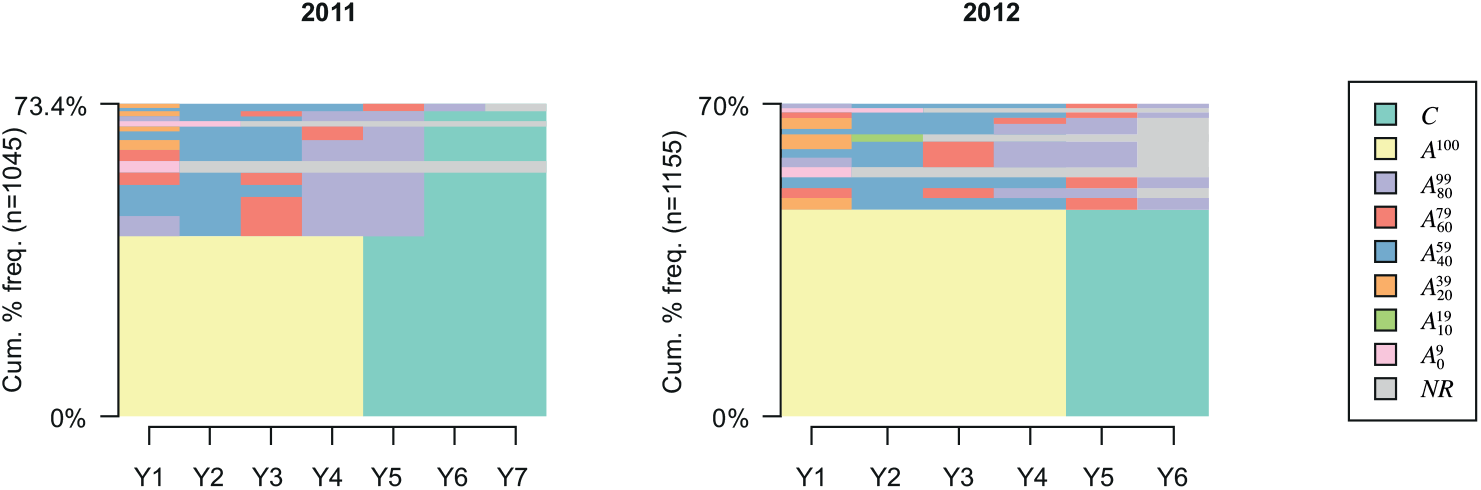
Frequency of academic progress trajectories in the C-2010 program, 2011 and 2012 student cohorts, UNAM Faculty of Medicine (n=2,200)

The 1996 class had an atypical behavior as mentioned previously, probably due to the prolonged university strike in 1999. There was a proportion of students in this class that showed a regular behavior the first three years, and in the fourth year there was an arrest that created a debt of up to 20% of curricular credits. Due to the difficulty of establishing a criterion to determine the best or worst type of academic trajectories, we selected the trajectory of “regularity”, understood as the accumulation of academic credits by the students, according to the curriculum and time period since the student’s first registration.

In order to compare different trajectories, percent distributions of academic trajectories were calculated for each cohort. Table 6 shows these data for the 2004 cohort of the C-1994 curriculum. For example, the trajectory *A*^*100*^*/4-*C*/3* can be interpreted this way: the student was regular during the first four years of the program (covered the total required credits established in the curriculum) and in the 5th year she completed all the credits required to obtain the degree (C state), she remained in this state during the sixth and seventh years. In Mexico medical schools require an extra year of mandatory social service after completing all the curricular credits, in order to obtain the MD degree. Since the social service has no credits, it was not included in our analysis.

**Table 6.** The more common academic progress trajectories in the C-1994 program 2004 student cohort, UNAM Faculty of Medicine (n=925).

| Trajectory | Frequency | Percentage |
| --- | --- | --- |
| $A^{100}/4-C/3$ | 338 | 36.5 |
| $A_{40}^{59}/2-A_{60}^{79}/2-A_{80}^{99}/1-C/2$ | 83 | 9.0 |
| $A_{80}^{99}/1-A_{40}^{59}/1-A_{60}^{79}/2-A_{80}^{99}/1-C/2$ | 54 | 5.8 |
| $A_0^9/1-NR/6$ | 41 | 4.4 |
| $A_{60}^{79}/1-A_{40}^{59}/1-A_{60}^{79}/2-A_{80}^{99}/1-C/2$ | 37 | 4.0 |
| $A_{20}^{39}/1-A_{40}^{59}/1-A_{60}^{79}/2-A_{80}^{99}/1-C/2$ | 34 | 3.7 |
| $A_{40}^{59}/2-A_{60}^{79}/1-A_{40}^{59}/1-A_{60}^{79}/1-A_{80}^{99}/1-C/1$ | 23 | 2.5 |
| $A_0^9/2-NR/5$ | 21 | 2.3 |
| $A^{100}/1-R2/1-A_{60}^{79}/2-A_{80}^{99}/1-C/2$ | 12 | 1.3 |
| $A_0^9/1-A_{40}^{59}/1-A_{60}^{79}/2-A_{80}^{99}/1-C/2$ | 10 | 1.1 |
| $A_{40}^{59}/2-A_{60}^{79}/2-A_{80}^{99}/2-C/1$ | 9 | 1.0 |
| $A_{40}^{59}/4-A_{60}^{79}/1-A_{80}^{99}/1-C/1$ | 9 | 1.0 |
| $A_{20}^{39}/1-A_{40}^{59}/1-A_{60}^{79}/1-A_{40}^{59}/1-A_{60}^{79}/1-A_{80}^{99}/1-C/1$ | 8 | 0.9 |
| $A_{20}^{39}/1-A_{10}^{19}/1-NR/5$ | 8 | 0.9 |
| $A_{20}^{39}/1-A_{40}^{59}/3-A_{60}^{79}/1-A_{80}^{99}/1-C/1$ | 7 | 0.8 |

Other prominent trajectories are those of the students that did not register for the following cycle (NR, not registered), for example the trajectory 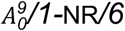(Table 6), where the student initially had a progress of less than 10% (delay of more than 90%, 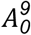), and in the next six years of follow-up he did not register courses (NR). This group represents 4.4% of the cohort. The 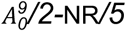 trajectory students, which were 2.3% of the cohort, had two years with a progress of less than 10%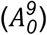 in the first and second year, and did not register (NR) in the following five years. The 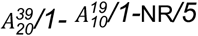 trajectory group, 0.9% of the student cohort, had a progress of 20% 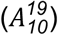 in the first year, in the second year of less than 10%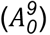, and in the following five years they did not register (NR). A total of 7.6% of the 2004 student cohort apparently abandoned the program during first or second year (grey band in Fig. 3).

In synthesis, regarding the 2004 cohort trajectories, it can be pointed out that 36.5% of the students had a regular and consistent route, and that 7.6% of the students left the program in the first or second year. In the first year 41.5% of the students had a regular trajectory, and in the fourth year this percentage decreased to 37.3%, as students advance in the curriculum 4.2% do not manage to pass the required courses.

Fig. 4 shows the student trajectories of the 2011 and 2012 cohorts that had the C-2010 curriculum. In these cohorts there is a larger proportion of students that had a regular trajectory (yellow and green portion of the graphs), which was higher than those of the C-1994 curriculum cohorts. It is not possible to state that this change in pattern of trajectories’ behavior was due to the different curriculum, since curricular change is a complex and multidimensional phenomenon, but we think it is possible to establish metrics that allow comparison of student cohorts that undergo different programs in different times. This metric can be regularity.

Table 7 shows data for the most common trajectories in the 2011 cohort of students (C-2010 program). The most frequent trajectory is the regular one, with 440 students (42.1%) of the total.

**Table 7.** The more common academic progress trajectories in the C-2010 program 2011 student cohort, UNAM Faculty of Medicine (n=1045).

| **Trajectory** | **Frequency** | **Percentage** |
| --- | --- | --- |
| $A^{100}/4-C/3$ | 440 | 42.1 |
| $A_{80}^{99}/1-A_{20}^{39}/1-A_{60}^{79}/1-A_{80}^{99}/2-C/2$ | 51 | 4.9 |
| $A_{20}^{39}/2-A_{60}^{79}/1-A_{80}^{99}/2-C/2$ | 46 | 4.4 |
| $A_{20}^{39}/3-A_{80}^{99}/2-C/2$ | 32 | 3.1 |
| $A_{60}^{79}/1-A_{20}^{39}/1-A_{60}^{79}/1-A_{80}^{99}/2-C/2$ | 29 | 2.8 |
| $A_0^9/1-NR/6$ | 29 | 2.8 |
| $A_{60}^{79}/1-A_{20}^{39}/2-A_{80}^{99}/2-C/2$ | 28 | 2.7 |
| $A_{20}^{39}/1-A_{20}^{39}/2-A_{80}^{99}/2-C/2$ | 23 | 2.2 |
| $A_{20}^{39}/3-A_{60}^{79}/1-A_{80}^{99}/1-C/2$ | 21 | 2.0 |
| $A_{20}^{39}/1-A_{20}^{39}/2-A_{60}^{79}/1-A_{80}^{99}/1-C/2$ | 13 | 1.2 |
| $A_0^9/2-NR/5$ | 13 | 1.2 |
| $A_{80}^{99}/1-A_{20}^{39}/2-A_{80}^{99}/2-C/2$ | 12 | 1.2 |
| $A_{20}^{39}/1-A_{20}^{39}/1-A_{60}^{79}/1-A_{80}^{99}/2-C/2$ | 12 | 1.2 |
| $A_{20}^{39}/4-A_{60}^{79}/1-A_{80}^{99}/1-NR/1$ | 9 | 0.9 |
| $A_{20}^{39}/1-A_{20}^{39}/3-A_{60}^{79}/1-A_{80}^{99}/1-NR/1$ | 9 | 0.9 |

Comparing the 2004 with the 2011 classes, a higher percentage of students completed the required credits in the 2011 class: 46.2% vs 41.5% in the first year of the program, 44.2% vs 39.0% in the second, 43.3 vs 37.6% in the third, and 42.9% vs 37.3 in the fourth. Comparing the percentage of students that developed a regular trajectory during the whole program, only 36.5% of the 2004 class (C-1994) had this trajectory, whereas the 2011 class (C-2010) 42.1% of the students developed this type of trajectory, a 5.6% difference. These two indicators, the percentage of advance in the regular state each year, and the percentage of students with regular trajectory, can account for the way students behave when fulfilling compliance with the program of studies. Changes in curriculum should be reflected in the way students move through the program.

There was a total of 16 generations that received the C-1994 curriculum, but we analyzed only five, to compare them with two generations that received the C-2010 program. The inclusion of the 2004 and 2005 cohorts had two reasons: the seven-year observation period, which did not overlap with the change of curriculum in 2010; and the proximity to the moment of curricular change (Fig. 5). This proximity is relevant due to the changes that occur in a school’s environment and context, the circumstances that surround the students were different in 1994 when compared to 2004 and 2005, even though it is the same curriculum.

**Figure 5.**
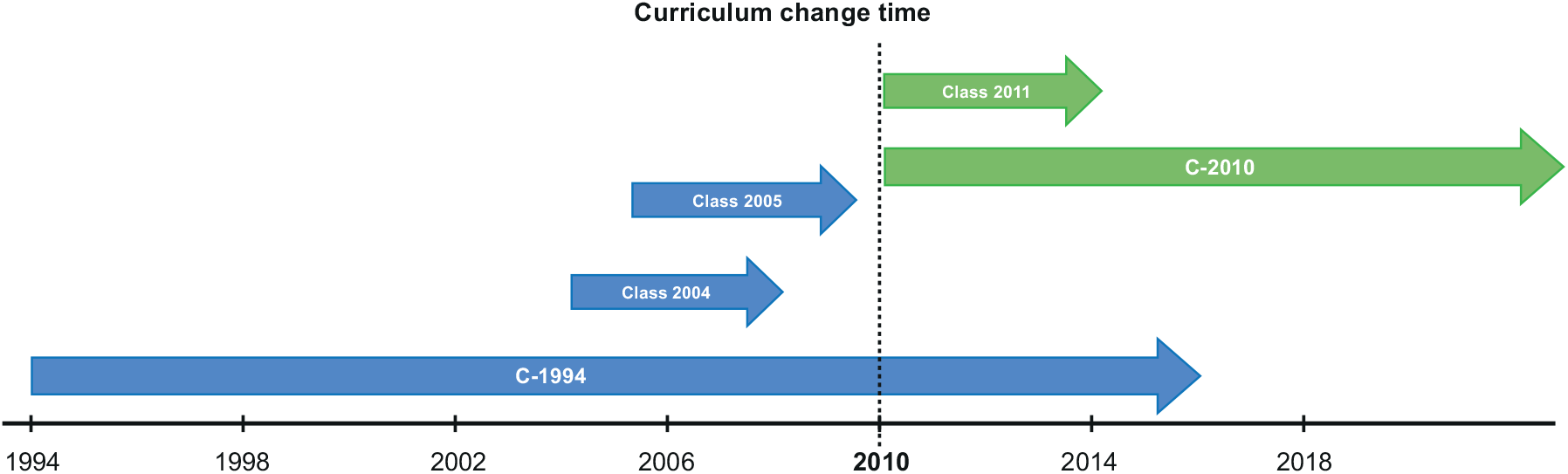
Timing of curricular change, UNAM Faculty of Medicine.

We compared the percentage of students that were in the regular state (*A*^*100*^) each year for the 2004, 2005 and 2011 classes (Fig. 6). The 2011 class with the newer curriculum had a higher percentage of students in the regular state each year, compared with the previous generations. If we had compared these generations with the traditional terminal efficiency parameter, we wouldn’t have been able to observe these longitudinal changes, since in the 6th year the difference is minimal (2%).

**Figure 6.**
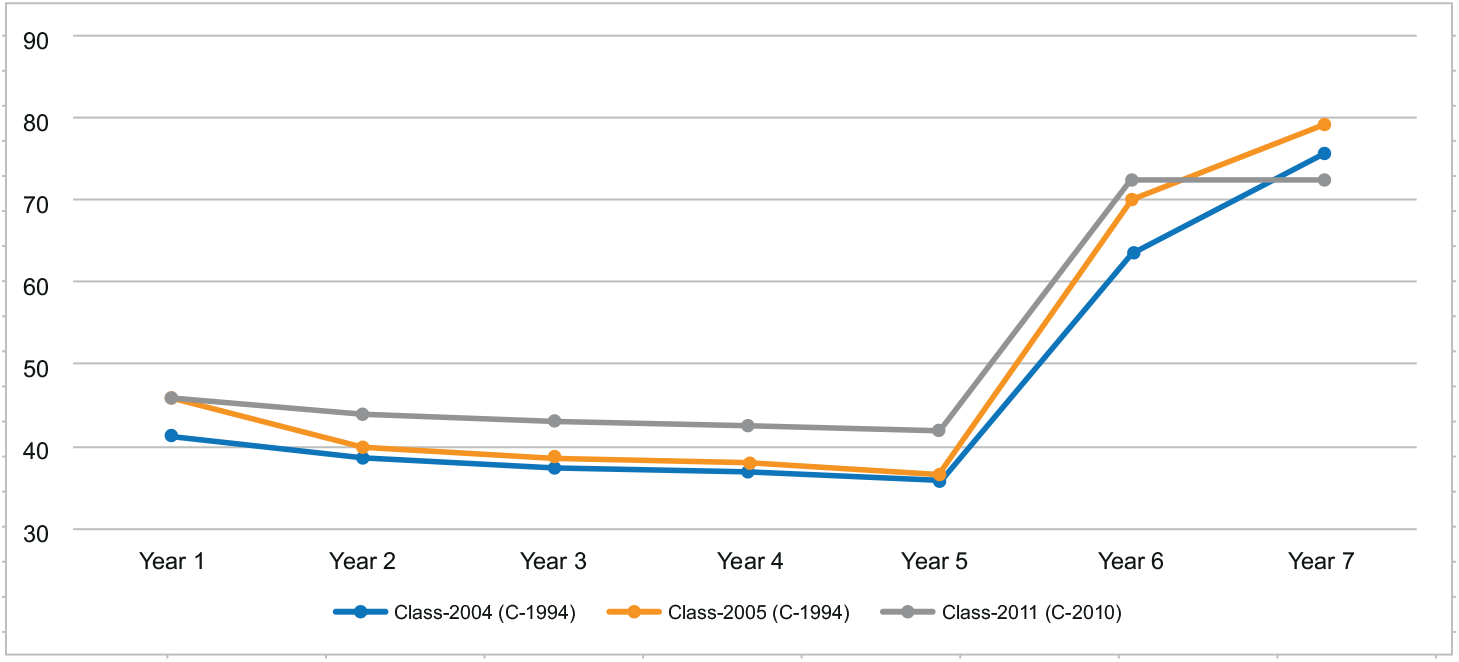
Comparison of student percentages in the “Regular” state (*A*^*100*^) in classes 2004 (C-1994), 2005 (C-1994) and 2011 (C-2010).

Figure 7 shows the percentage of students that developed a regular trajectory (*A*^*100*^) throughout the program, for the 1994, 1995, 2004, 2005, 2011 and 2012 classes (the last two had the C-2010 curriculum). The percentage of students with a regular trajectory is higher in the C-2010 cohorts by about 10%. If the tendency shown in the graph is maintained, it would be expected that in a manner similar to the C-1994 curriculum, where the first generation had 28.3% of students with a regular trajectory and in the second cohort the percentage leveled off at about 36%, the C-2010 would be stabilized after the 2012 class at about 46%.

**Figure 7.**
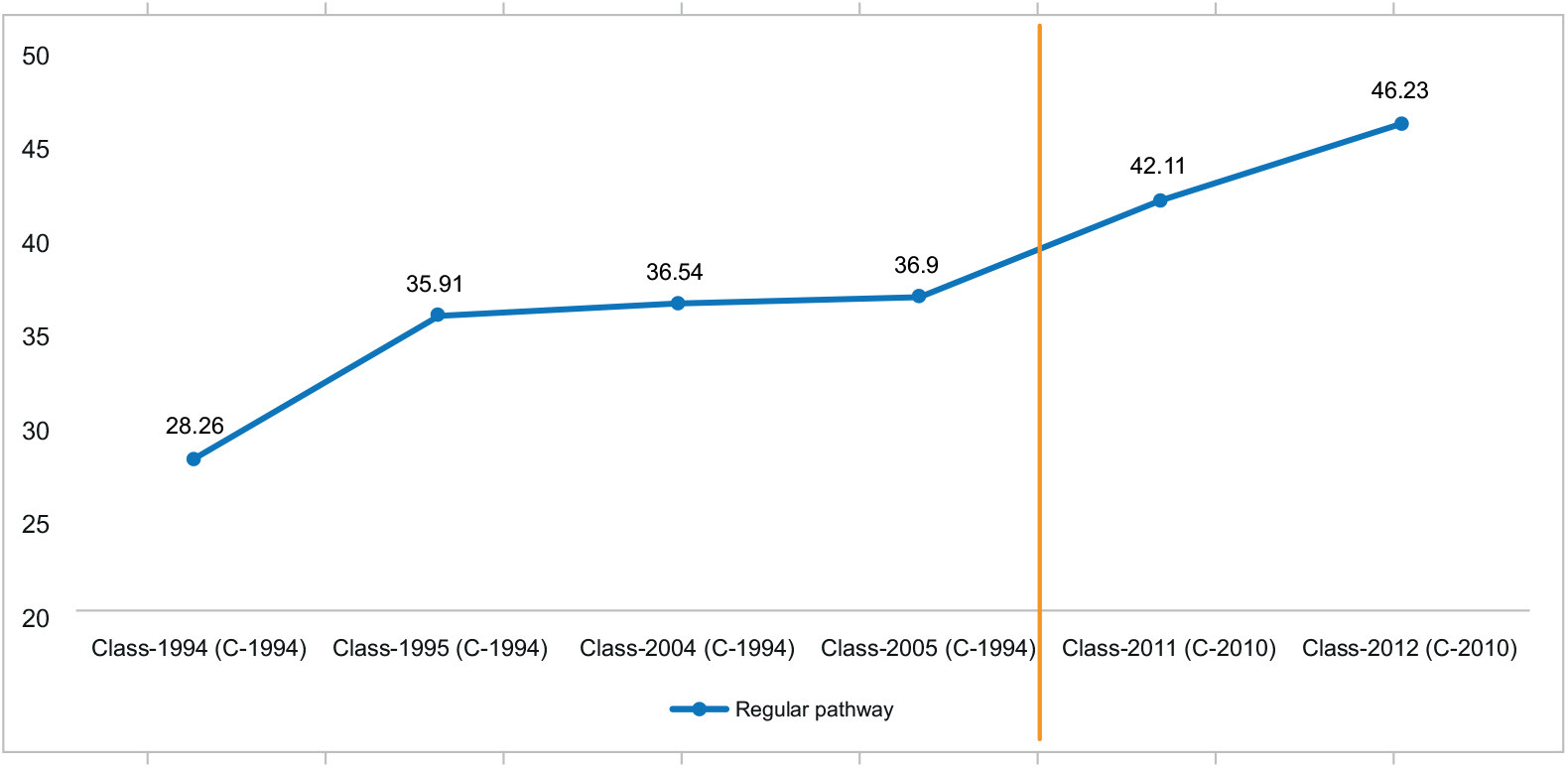
Comparison of the percentage of students with regular trajectory (*A*^*100*^) by class and curriculum.

## Discussion

Medical education scholars have published extensively about the importance of curriculum and program evaluation, emphasizing the need for developing these evaluative activities in a professional manner [65,66]. The proposed methodology can help curriculum evaluation teams that collect information relevant to program evaluation, to add these results to one of the most frequently used program evaluation methods, the CIPP model (Context, Input, Process and Product), that allows evaluation of the implementation process and its outcomes [67].

The Accreditation Council for Graduate Medical Education (ACGME) from the United States of America, defines program evaluation as “the systematic collection and analysis of information related to the design, implementation, and outcomes of a program, for the purpose of monitoring and improving the quality and effectiveness of the program” [68]. The main purpose of this type of evaluation is to identify sources of variation in curriculum outcomes, and to establish if these variations are desirable or not. Evaluation models have been predominantly focused in measuring only curricular outcomes, however in more recent times the new models of evaluation are concentrating on the dynamics of the implementation process that allow proposal of improvements [67]. We selected an evaluation strategy that is theoretically grounded, to illustrate the use of the method in a medical education case study.

The CIPP evaluation model has four dimensions that taken together allow evaluators to propose systemic improvements. The first three dimensions (Context, Input and Process) are targeted to improve the educational process, whereas the fourth dimension (Product) is more focused on summative evaluations [66]. The evaluation of Process in the CIPP model is frequently utilized during the implementation of a program and can provide information about the formative activities. In a curriculum that operates in complexity, as is the case in medical schools, attention to process can generate useful data to manage and innovate the program. We recover the trajectories of medical students to account for the process dimension of the curriculum, with the goal of showing the students’ academic advance in two different curricula, and to build a measure that helps provide a broader view of the formative process. This measure is regularity, which allows the comparative visualization of the students’ transit through two different medical school programs.

This paper proposes a method of analysis that accounts mainly for the Process and Product dimensions, where the analysis of students’ academic pathways provide information about their states and formative transit through the programs, following their academic histories, accounting for the obtained credits (as well as the events of dropouts, delays or reinsertions in the process), and showing the obtained results or Products.

### The curriculum change phenomenon

The straightforward measurement of results generated by complex phenomena is an ever-present need in higher education, required to promote reflection and support decision making even though there is always theoretical and epistemic uncertainty in these processes [69]. Medical education can be viewed as an interlinked system of complex subsystems [53], where major curricular changes are frequent phenomena [6]. The measure of educational changes is a difficult task, and there is a necessity for an objective global measure to evaluate its effects in the student population.

We propose a metric, the regularity of academic progress throughout a time period, built through a process of identifying academic trajectories in the regular category. This is based on the assumption that the behavior of academic trajectories synthesizes the results derived from curricular changes in a higher education program. Changes in a curriculum are subject to diverse forces, among its more visible components are the following: participants’ resistance to change, the assumptions and models of curriculum design, faculty development, university regulations, social context and local circumstances.

### The method to identify student academic trajectories

The proposed method to identify student academic trajectories requires the standardization of data in an objective fashion, which in this particular study was done retrospectively. This methodology does not integrate qualitative data, which are difficult to mix with the quantitative data used in our study. This does not mean that qualitative information is less important to achieve and maintain curricular changes, and that should be the object of further study [19]. The concept of student academic trajectories is traditionally applied to studies that use sociodemographic variables, at the local or regional levels [39]. Its role in the analysis of correlations of changes in academic trajectories and curricular modifications has not been well studied [22].

Our method proposes as the basic unit of measurement the number of credits per course that is established by the curricular plans under study. This measurement unit allowed the building of each transition state and academic trajectory. One of our assumptions was that the number of study hours dedicated to each course has a correlation with the level of course difficulty. This assumption can generate bias, since the degree of difficulty of each course is not linear, but is a function of the students’ background, metacognitive abilities, acquired knowledge, and perseverance [43,53]. Our methodology and the available information allowed us to quantify each and every one of the students’ academic trajectories, although due to the study goals we present only the 15 more common trajectories. This means that the trajectories’ singularity was not analyzed in this study, which opens the possibility of further curricular analyses.

The uncertainty involved in the correlation between alterations in academic trajectories and the curricular changes, was attenuated by the temporal proximity and the condition of mutual exclusion between programs. Temporal proximity allows the assumption that environmental conditions among the cohorts were similar, with respect to economic, technological and political factors. The condition of mutual exclusion between curricula means that the students were subjected to one and only one curriculum, although there was a time period during which both programs were active. Regularity is a quantitative metric, which eventually requires the integration of qualitative data. This would make it possible to generate systemic judgment values about the production of medical education spaces [70].

### Student population

In our study, the analysis of academic trajectories was applied to cohorts of medical students that were subject to different educational programs at a large public medical school in Mexico. The institution had the challenge of implementing a competency-based educational model, improving the integration of the program contents with a focus on professionalism, and faculty development activities. Five student cohorts of a traditional curriculum (C-1994) were analyzed as well as the first two generations of a new program (C-2010). The follow-up involved the quantification of the advance in credits for each student at the end of each academic cycle, in relation to the fixed required time period to cover the courses. With these data, groups of academic advance were identified which we labeled transition states. This allowed us to build patterns of academic performance of students throughout the program. The comparison between the different curricula was focused on the students that were always regular and graduated in the time period established in the program. Two additional years were considered to observe the trajectories’ behavior posterior to the time established in the curriculum.

### Variations in student academic trajectories

The trajectories of the cohorts in the older plan show two patterns of regularity: in the 1994-1996 generations regularity decreases while students advance in the program, while in the newer plan, the 2004 and 2005 cohorts regularity is constant. Evidence from several administrative changes that the university underwent in these periods, suggests that the populations of students are similar and that the differences in academic trajectories can be attributed, at least in part, to the different curricula. In both programs about 80% of the students finished their studies by eight years after admission, although there was a 5% higher rate of regular trajectory in the C-2010 curriculum compared to the previous (C-1994) program. The structure of both plans does not allow students to advance if they do not pass a course, so they cannot catch until they pass the required courses for that year. The first year of the program is crucial in order to have a regular trajectory from the beginning, which determines the subsequent trajectories. The challenges and failure of many students during the first year of college evidence the limitations of the high-school graduates in our educational system [11,36]. High-school curricula and graduation profiles should be articulated with college admission requirements and profiles [12,39].

The differences in academic trajectories can be due to multiple factors: the curricular integration of contents in the newer curriculum, improved students’ learning and studying techniques, faculty development initiatives, different academic practices, use of technology and simulators [18,25]. These activities and many others can influence academic performance in medical students, and these should be studied in higher detail in subsequent studies.

Faculty development is one of the major factors that can determine faculty performance and student learning [24,25]. Teacher training and professional development influences their teaching methods and beliefs in student learning, although it’s difficult to move people from their comfort zones. Organizational changes and incentives can have an important effect in faculty [12]. We believe that more research should be done about curricular changes in higher education, and that the methodology described in this paper can be useful to evaluate the changes that occur in students’ academic trajectories. The results of similar studies can be included in program evaluation designs to supplement the data and information obtained with more traditional sources [16,29].

Our study has some limitations: we analyzed only the transit of students in a single medical school in an emerging economy country, so generalization of our results should be done with caution. We did not include individual student socioeconomic variables that can influence academic performance, and do not have posterior follow-up of the students after graduation and their achievement in the healthcare system. Even though the methodology can be applied theoretically to any field of knowledge, we studied only medicine, so the findings have to be replicated in other areas of the professions. UNAM Faculty of Medicine is one of the largest medical schools in the world, and the amount of data used in the analysis is large enough to identify diverse trajectories, in smaller schools the methodology could be less applicable. On the other hand, the methodology used in this study has not been reported, as far as we know, in the area of curriculum change and program evaluation, and there are very few studies evaluating major curricular changes in medical schools with data from before and after the modification.

## Conclusions

The proposed method allows the construction of academic trajectories to analyze, separate, abstract and quantitatively synthesize the alterations that occur with curricular modifications.

The analysis of students’ academic pathways offers valuable information for the evaluation of curricular changes. This information is difficult to obtain with more traditional cross-sectional studies. The study does not provide proof of causality regarding the educational impact of different programs, but it can be useful to complement the array of program evaluation strategies in medical education.

## Data Availability

Data are available on request from the authors

## Acknowledgments

The authors would like to thank the academic personnel of the Coordination of Educational Development and Curricular Innovation (CODEIC) and the Coordination of Open University and Distance Education (CUAED), both from the National Autonomous University of Mexico, for their support for this study.

## S1 Appendix

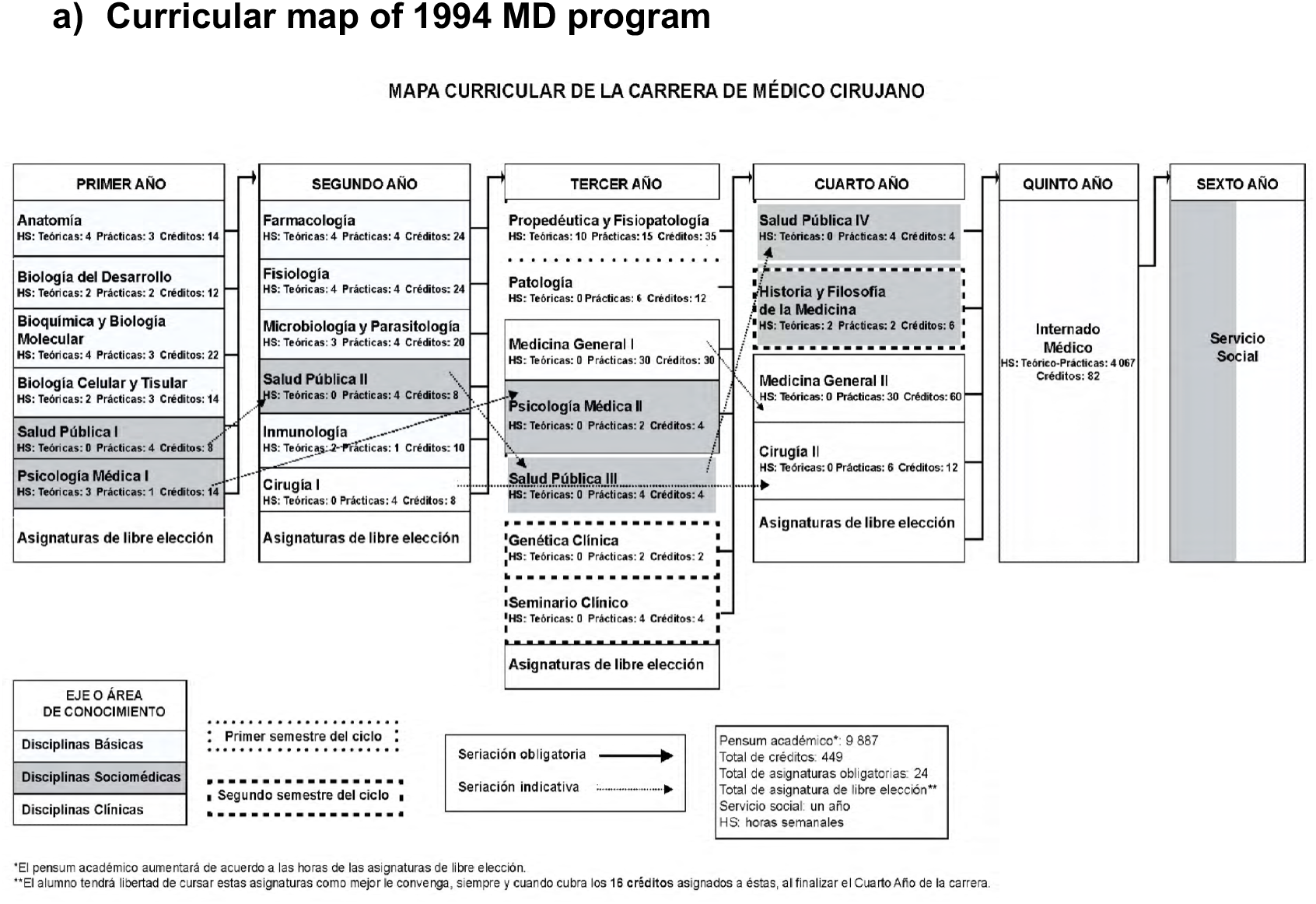
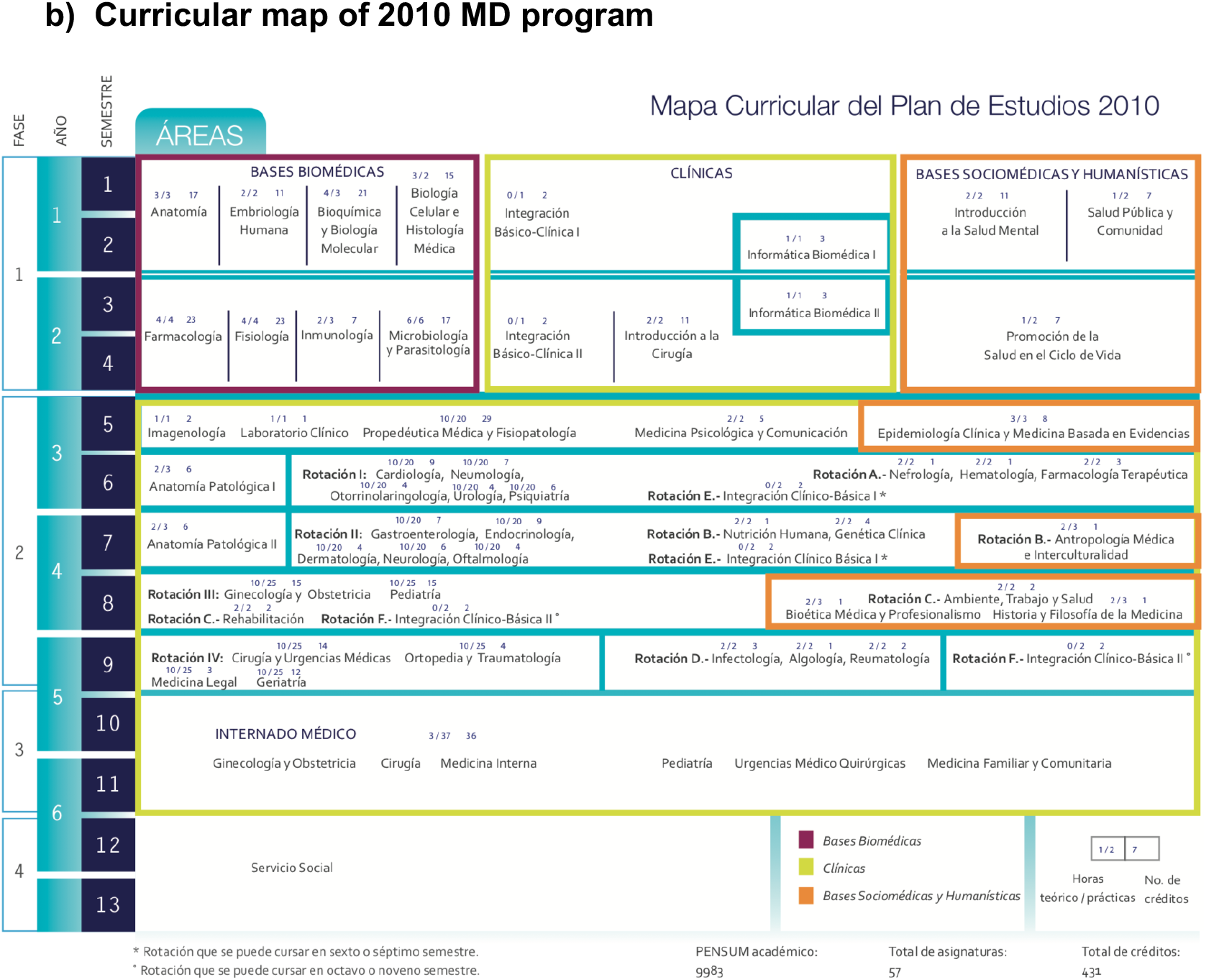

## S2 Appendix

**Comparative table of the general characteristics of the 1994 MD Curriculum and the 2010 program**.

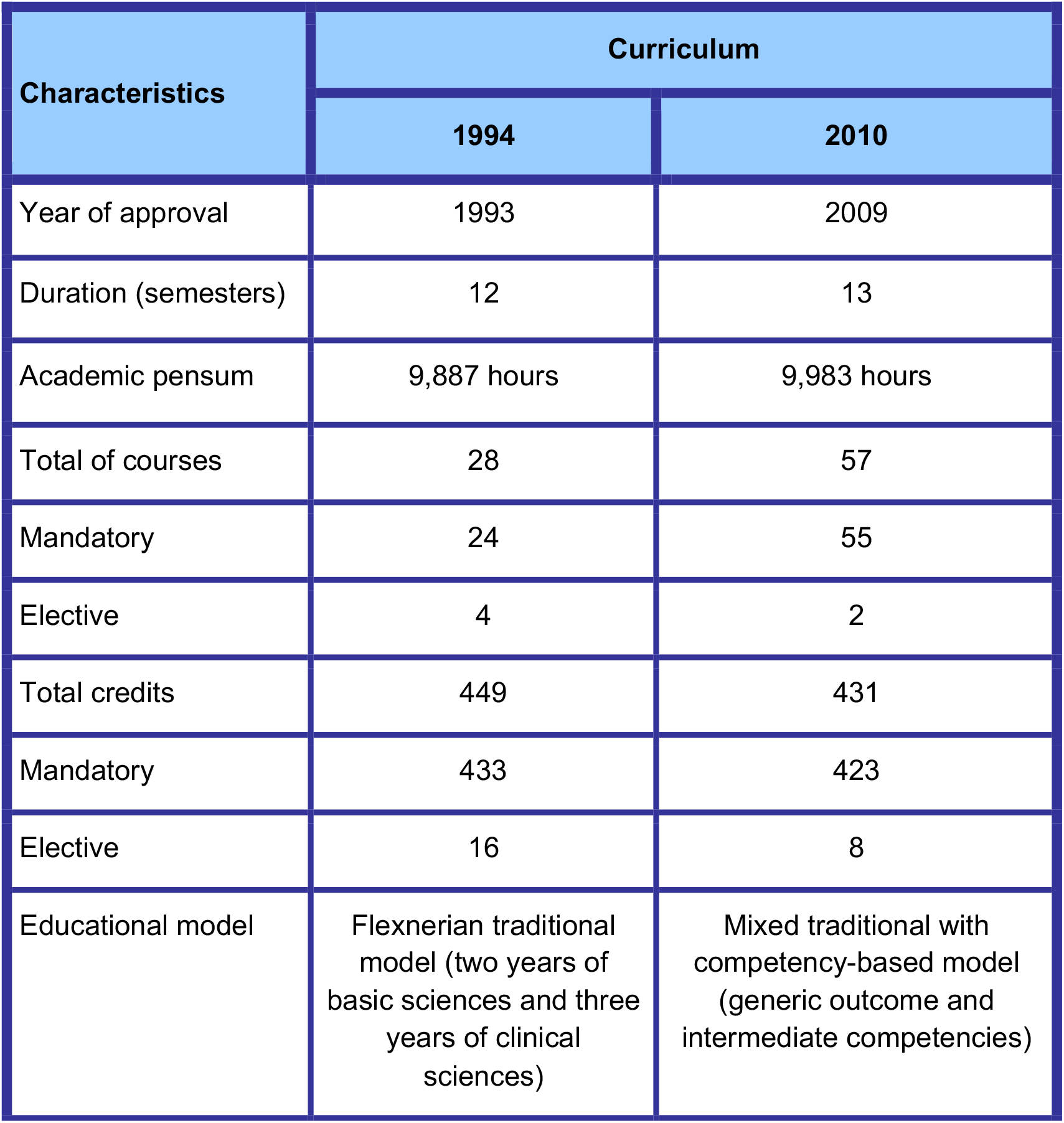
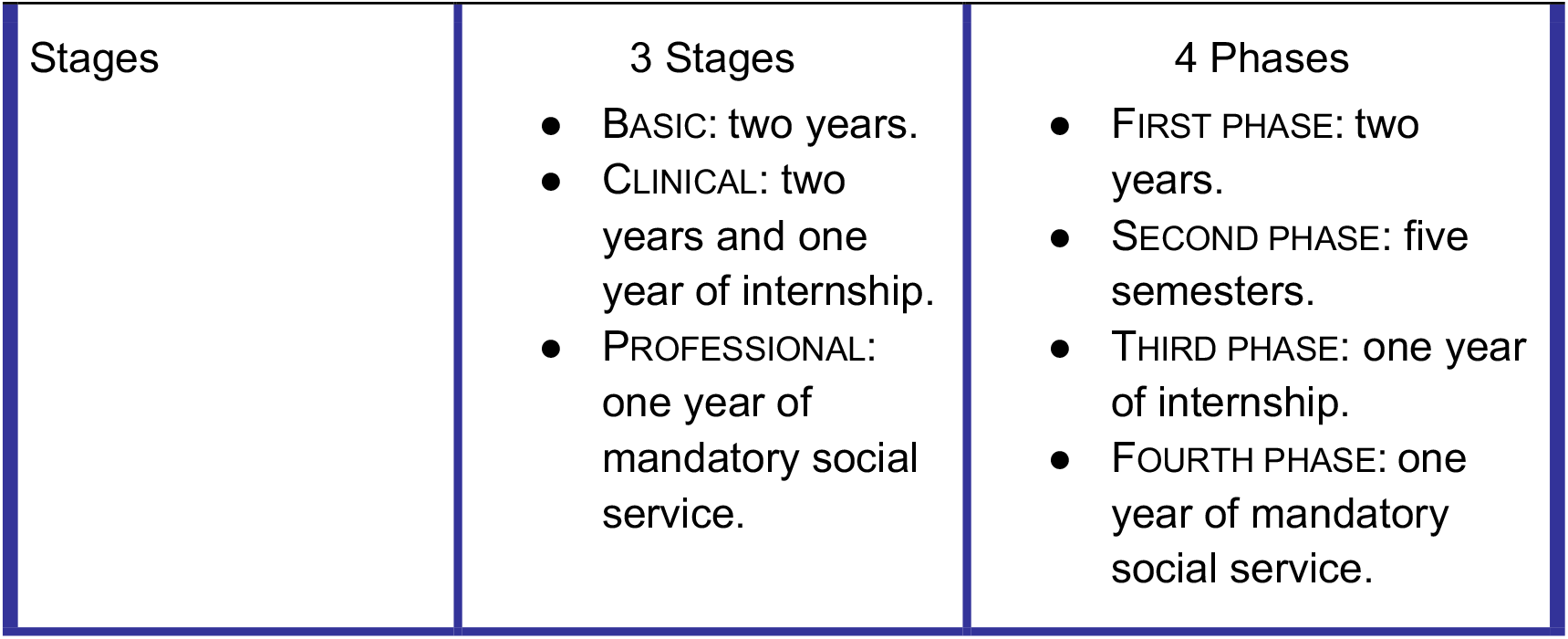

